# Systematic evaluation of SARS-CoV-2 spike protein derived peptides for diagnosis of COVID-19 patients

**DOI:** 10.1101/2020.09.01.20186387

**Authors:** Yang Li, Dan-yun Lai, Qing Lei, Zhao-wei Xu, Hongyan Hou, Lingyun Chen, Jiaoxiang Wu, Yan Ren, Ming-liang Ma, Bo Zhang, Hong Chen, Caizheng Yu, Jun-biao Xue, Yun-xiao Zheng, Xue-ning Wang, He-wei Jiang, Hai-nan Zhang, Huan Qi, Shu-juan Guo, Yandi Zhang, Xiaosong Lin, Zongjie Yao, Pengfei Pang, Dawei Shi, Wei Wang, Xiao Yang, Jie Zhou, Huiming Sheng, Ziyong Sun, Hong Shan, Feng Wang, Xionglin Fan, Sheng-ce Tao

**Author notes:** These authors contribute equally to this work (Yang Li, Dan-yun Lai, Qing Lei, Zhao-wei Xu, Hongyan Hou). Corresponding Authors, (S.-C. Tao), (X.-L. Fan), (F. W.) and (H. S.).

## Abstract

Serological test plays an essential role in monitoring and combating COVID-19 pandemic. Recombinant spike protein (S protein), especially S1 protein is one of the major reagents for serological tests. However, the high cost in production of S protein, and the possible cross-reactivity with other human coronaviruses poses unneglectable challenges. Taking advantage of a peptide microarray of full spike protein coverage, we analyzed 2,434 sera from 858 COVID-19 patients, sera from 63 asymptomatic patients and 610 controls collected from multiple clinical centers. Based on the results of the peptide microarray, we identified several S protein derived 12-mer peptides that have high diagnosis performance. Particularly, for monitoring IgG response, one peptide (aa 1148-1159 or S2-78) has a comparable sensitivity (95.5%, 95% CI 93.7-96.9%) and specificity (96.7%, 95% CI 94.8-98.0%) to that of S1 protein for detection of both COVID-19 patients and asymptomatic infections. Furthermore, the performance of S2-78 IgG for diagnosis was successfully validated by ELISA with an independent sample cohort. By combining S2-78/ S1 with other peptides, a two-step strategy was proposed to ensure both the sensitivity and specificity of S protein based serological assay. The peptide/s identified in this study could be applied independently or in combination with S1 protein for accurate, affordable, and accessible COVID-19 diagnosis.

**One Sentence Summary:** Eight S protein-derived peptides, particularly S2-78 (aa 1148-1159), are of high performance for diagnosis of COVID-19 as well as discrimination of other coronaviruses.

---

COVID-19 is caused by SARS-CoV-2[1,2] and is a global pandemic. By August 7, 2020, 18,982,658 cases were diagnosed and 712,266 lives were claimed (https://coronavirus.jhu.edu/map.html)[3]. To put the pandemic under control, one of the essential options is to perform fast, reliable and affordable diagnosis. Although nucleic acid test (NAT) is the reference standard for diagnosing COVID-19 with high sensitivity and accuracy, however, false negative results were commonly observed[4,5]. The immunological/ serological test, for example, monitoring the SARS-CoV-2 specific IgG and IgM responses, provides important information to improve the accuracy of diagnosis[4,5]. In addition, serological test is suitable for population screening in high risk regions or among close-contact people, as well as surveillance of the pandemic spreading or assess the infection rate of general population[6-8]. Moreover, antibody response is reported to associated with disease severity or clinical outcomes[9,10].

S protein is the preferential antigen for serological assay. The key reagent of the S protein based serological assay is the recombinant protein. However, the production of the S protein is difficult and costly[11]. The inconsistency among different manufacturers or even batches might contribute to the variability of commercial assays with the same antigen[7,12]. Limited production capacity and high cost of recombinant protein preparation is the bottleneck, particularly for remote regions or poor countries. In addition, it should be concerned that the cross-reactivity of infections of other human coronaviruses may cause false positive results, especially for those four common cold causing coronaviruses, *i.e*., HCoV-OC43, HKU1, NL63 and 229E, which are circulating in population[4,11,13]. It was reported that S1, compared with full length S protein, exhibit less cross-reactivity due to the less similarity of S1 subunit among the human coronaviruses than that of S2[4]. To develop highly specific serological test, more efforts are needed to identify sections of S protein that are highly immunogenic and less homologous to other related coronaviruses[6,11].

Spike protein derived peptides that can elicit antibodies in COVID-19 patients has been reported in several studies[14-16], including one of our previous work on epitope mapping with a small sample cohort[17]. For instance, antibody against S2-78 (aa 1148-1159) and S2-22 (aa 812-823) have high positive rates in COVID-19 patients. However, whether those peptides are suitable for diagnosis is still unknown. Herein, to fully evaluate the diagnostic value of the S protein derived peptides, a total of four cohorts of sera, consisted of 2,434 sera from 858 COVID-19 patients, sera from 63 asymptomatic patients and 610 controls were used. Eight peptides were verified to have high potential for diagnosis, particularly, one peptide, S2-78 has a comparable diagnosis performance as that of S1 protein for COVID-19 patients and asymptomatic infections. By combining S2-78 IgG/ S1 IgG with other peptide/s, we purposed a two-step strategy that can ensure both sensitive and specific diagnosis for COVID-19.

## Results

### Four independent cohorts of samples were collected and designed

To fully evaluate the diagnostic potential of the spike derived peptides, sera from four cohorts of COVID-19 patients and controls from multiple medical centers of China were collected (**Table 1**). 1) Cohort 1 consists of 55 sera from convalescent COVID-19 patients and 18 controls[17]; 2) Cohort 2 includes 2,360 sera from 784 in-hospital COVID-19 patients and 542 sera from a variety of controls. To accurately evaluate the peptides for diagnosis, for each patient, one serum sample that collected at least 21 days after onset of symptoms was selected according to the suggestion of WHO (World Health Organization) for antibody laboratory test[18]. As a result, in total, 729 sera were selected. The control groups include two types of samples. The first type is sera collected from hospitals, including sera from healthy people (n=92), upper respiratory infections (URI, n=104), patients with autoimmune diseases (AID, n=120), lung cancer patients (n=41) and patients with other diseases (n=112) that consist of cardiovascular or cerebrovascular diseases (34.2%), diabetes (9%), non-lung cancers (7.2%) and others. The second type is negative references provided by National Institutes for Food and Drug Control, including N11-N25 of National Reference Panel for 2019-nCoV (SARS-CoV-2) IgG Antibody Detection Kit (370096-202001) and other identified controls. 3) Cohort 3 includes 19 COVID-19 patients from another hospital and 50 healthy controls. 4) Cohort 4 consists of asymptomatic patients defined as positive either on NAT (nucleic acid test) or antibody test conducted by a commercial assay, and with a SARS-CoV-2 exposure history (see **methods**).

**Table 1.**
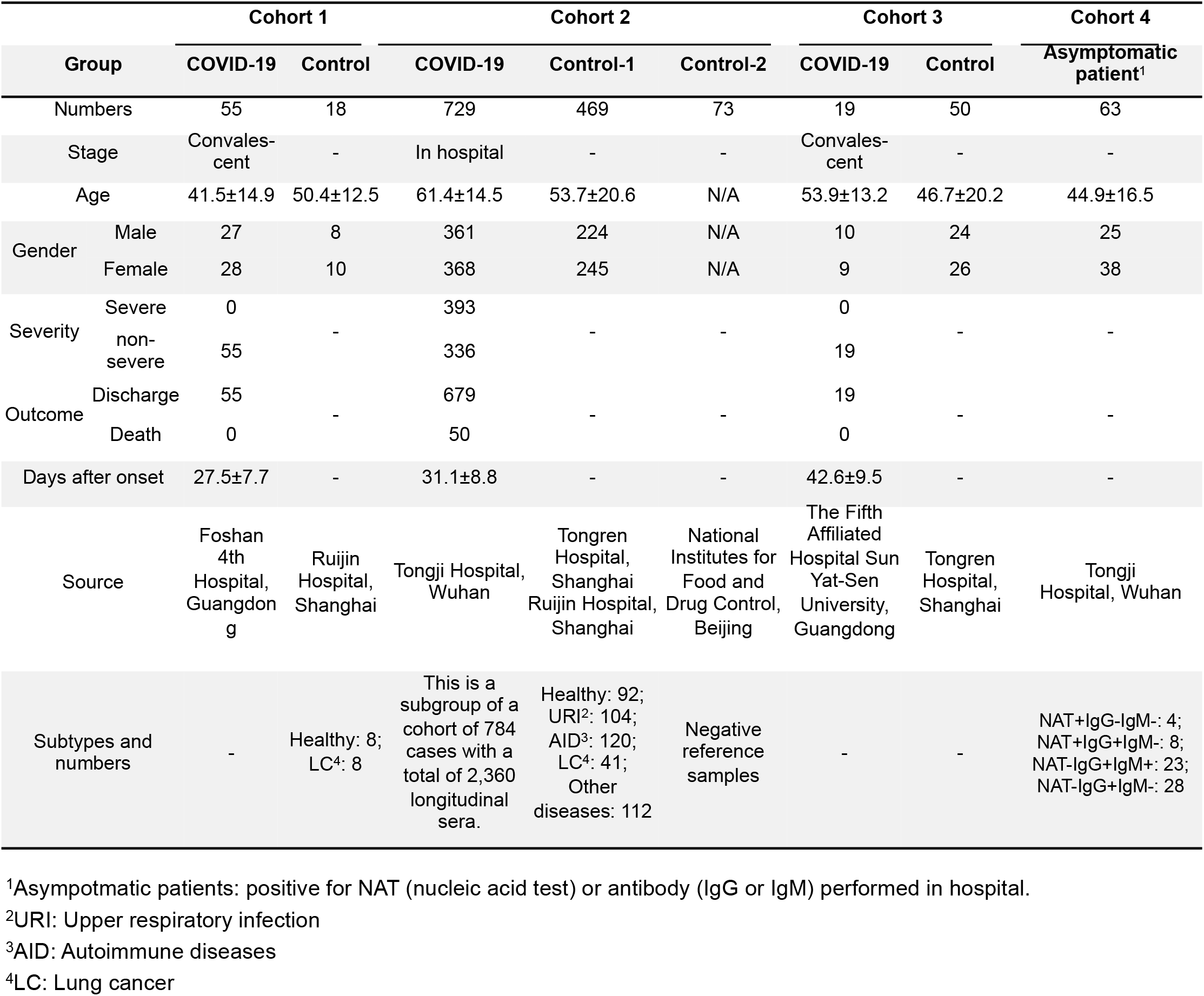
Patients and controls involved in this study.

### Peptide S2-78 and several other peptides exhibit high diagnostic values

To identify which section of S protein has diagnostic value, it is necessary to survey the entire protein on a systematic way. We took advantage of a previously constructed peptide microarray which has full coverage of S protein[17], analyzed 55 convalescent sera of COVID-19 patients along with 18 control sera (Cohort 1, **Table 1**). Overall, significant bindings of both IgG and IgM were observed in patient group, while the signals were low in the control group (**Fig. 1a**). Several peptides, *e. g*., S2-78 and S1-93 exhibit strong IgG antibody bindings and high response frequencies in patients, we define these peptides as “significant” peptides, these peptides may have diagnostic values (**Fig. 1b**). To test whether the peptide specific IgG antibody responses are concentration dependent, 3x serially diluted peptides (0.9, 0.3 and 0.1 mg/mL) were printed and immobilized on the microarray. As expected, the averaged signals of the patient group, but not the control group, are proportional to the concentrations of the peptides (**Fig. 1c**).

**Figure 1.**
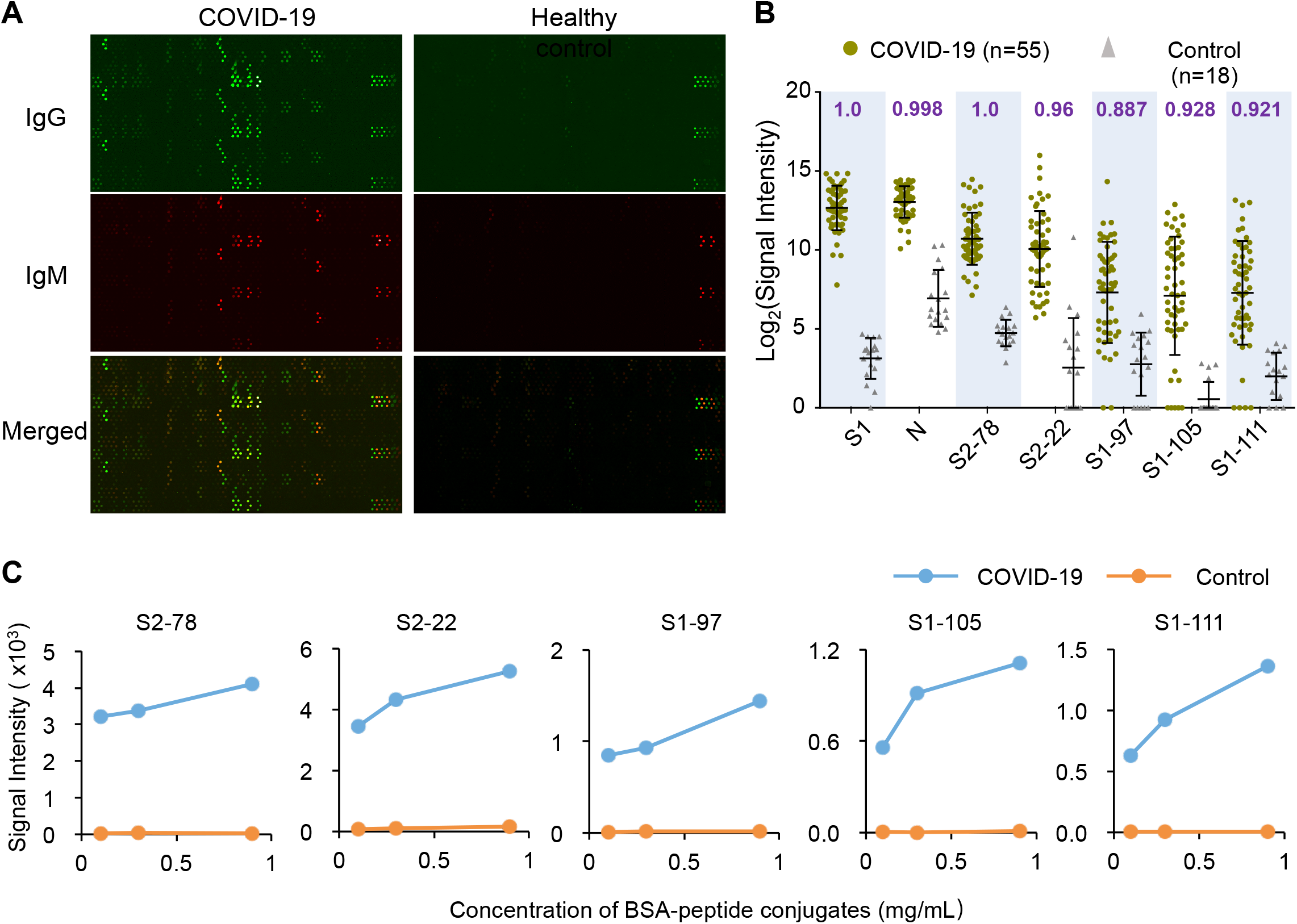
Peptides with strong IgG antibody responses were identified by Cohort 1. **A**. Representative images of peptide microarray profiled by sera from a COVID patient and a healthy control. IgG (green) and IgM (red) were detected simultaneously. **B**. Box plot of IgG antibody responses against SI protein, N protein and some significant peptides for COVID-19 patient group (n=55) and control group (n=18) of Cohort 1. Each spot indicates one serum sample. Data are presented as box plots where the middle line is the mean value, and the upper and lower hinges are mean values ± SD. AUC (area under curve) values are labeled for each peptide or protein on the top of the box plots. **C.** Averaged signal intensities of IgG antibody responses against the indicated peptides at different concentrations, *i.e*. 0.1, 0.3 and 0.9 mg/mL, both for COVID-19 patients (blue line) and control group (orange line).

To extensively evaluate the peptides for diagnostic application, a larger cohort (Cohort 2, **Table 1**) of samples were screened by a revised peptide microarray that contains only one peptide concentration (0.3 mg/mL) for high-throughput analysis. To ensure the data generated from different microarrays are comparable, we prepared a positive reference sample by pooling 50 randomly selected patient sera. This reference sample was then tested on all the microarrays for normalization (see **methods**). Consistent results were achieved for most of the peptides. AUC (area under curve) values of IgG or IgM for each peptide were calculated. Eight peptides, *i. e*., S2-78, S1-97, S1-93, S1-101, S1-111, S2-97, 51-105 and S2-22 are of high performance, the AUCs of IgG or IgM against these peptides for both Cohort 1 and 2 are above 0.85. Specifically, for Cohort 2 the AUC values with 95% CI (confidential intervals) for S2-78, S1-97, S1-93, S1-101, S1-111, 52-97, S1-105 and S2-22 were 0.99 (0.986-0.995), 0.954 (0.942-0.965), 0.934 (0.92-0.948), 0.932 (0.917-0.947), 0.929 (0.915-0.943), 0.922 (0.907-0.938), 0.909 (0.893-0.926), and 0.866 (0.846-0.886), respectively (**Fig. 2a**). For IgM, only S2-78 has a AUC >0.85, *i. e*., 0.953 (0.941-0.964) (**Fig. 2a**). As expected, IgG and IgM for both N protein and S1 protein exhibited high performance. We further examined the antibody responses in different groups for each peptide (**Fig. 2b-e, Fig. S1**). Consistently, signals for COVID-19 group are significantly higher than that of the negative samples in all groups. It is noted that the signal intensity for S1 IgG is generally higher than that of any single peptide in the group of COVID-19 patients, this may because there are multiple antibody binding sites on S1 protein. However, slightly higher signal is also observed in control group for protein antigens, demonstrating non-specific binding while it is largely eliminated for synthetic peptides. It is suggested that more sensitive detection platforms or higher antigen concentration might improve the performance of peptides for diagnosis.

**Figure 2.**
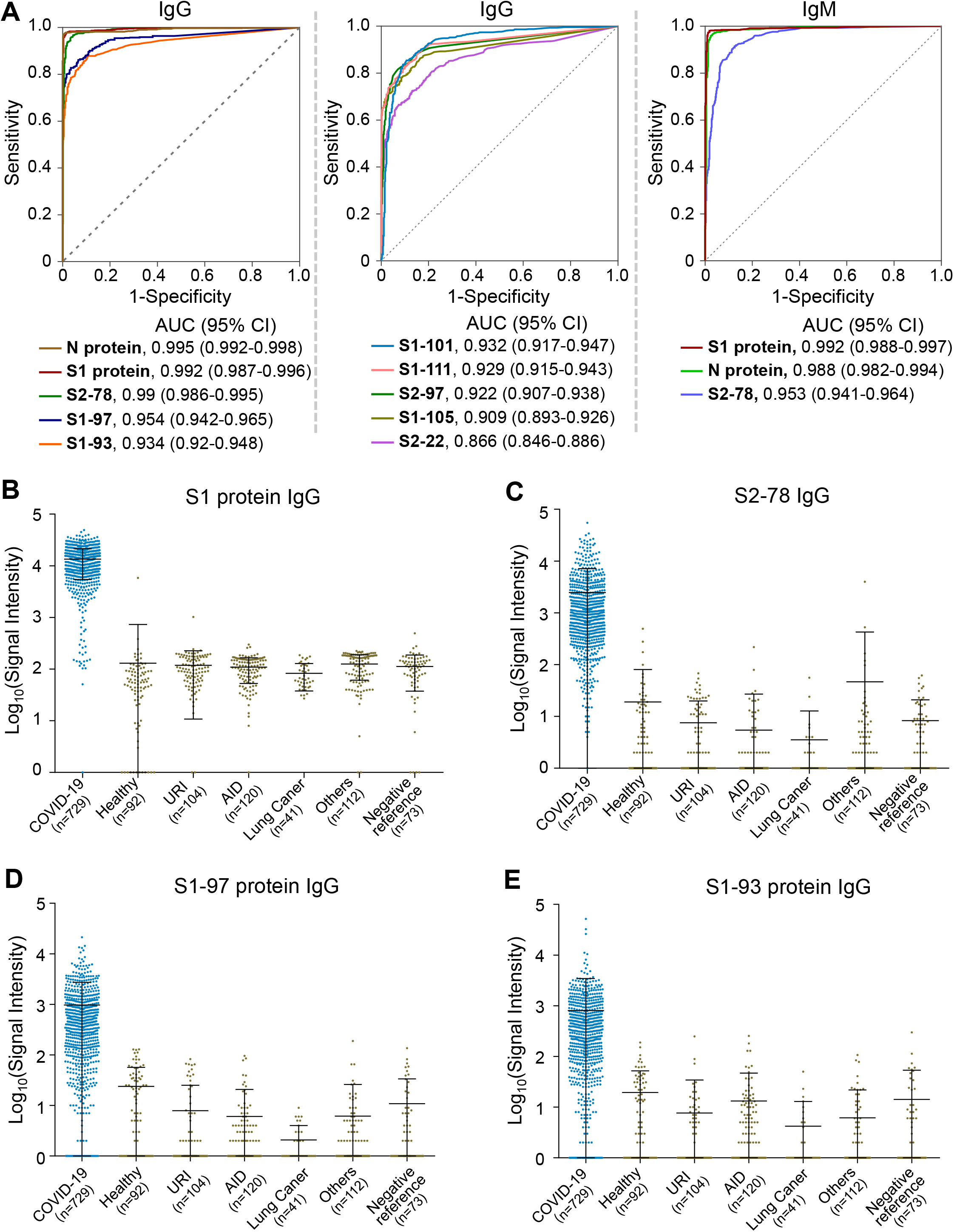
Evaluation significant peptides by Cohort 2. **A**. ROCs (receiver operating curves) of the peptides or proteins for discrimination of the COVID-19 group (n=729) and the control group (n=542). The AUC values with 95% Cl (confidential intervals) are provided for all the peptides and proteins. **B-E**. signal distributions of anti-S1 IgG (B), anti-S2-78 IgG (C), anti-S1-97 IgG (D) and anti-S1-93 (E) in COVID-19 patients (blue) and control groups (yellow). Sample size is indicated for each group. URI: upper respiratory infection, AID: patients with autoimmune diseases.

### The diagnostic performance of S2-78 IgG is comparable to that of S1 IgG for COVID-19 patients

We next focused on S2-78, the peptide of best performance for detection. Optimal Youden index of ROC (receiver operating characteristic) curve was used to set the cutoff value. The specificity, sensitivity and overall accuracy (95% CI) of S2-78 IgG for detection of COVID-19 are 96.7 (94.8-98.0%), 95.5% (93.7-96.9%) and 96% (94.8-97%), respectively, which are slightly lower than that of S1 IgG (**Table 2**). Since serological test is essential for population screening, we calculated the PPV (positive predict value) and NPV (negative predict value) of two assumed prevalence rates. One is 0.04 for general population originated from the situations of Wuhan, China[19] and Netherlands[20]. The other is 0.5 for a high risk population[7]. For prevalence rate of 0.5, both PPV and NPV of S2-78 IgG is similar to that of S1 IgG, however, for prevalence rate of 0.04, the PPV is only 54.7%, although the NPV is extremely high, suggesting the antibody detection of S2-78 could effectively exclude negative ones but may generate high false positive rate at low rate. However, for low prevalence rate, since the number of real positive are very low, it might be acceptable to perform additional test by using other antigens to improve the overall performance.

**Table 2.** The overall performance of S1 and S2-78 for diagnosis

|  |  | S1 |  | S2-78 |  |
| --- | --- | --- | --- | --- | --- |
|  |  | Positive | Negative | Positive | Negative |
| COVID-19 |  | 707 | 22 | 696 | 33 |
| Control |  | 2 | 540 | 18 | 524 |
| Specificity (95% CI) |  | 99.6% (98.7-100%) |  | 96.7% (94.8-98.0%) |  |
| Sensitivity (95% CI) |  | 97% (95.5-98.1%) |  | 95.5% (93.7-96.9%) |  |
| Accuracy (95% CI) |  | 98.1% (97.2-98.8%) |  | 96% (94.8-97%) |  |
| Prevalence |  |  |  |  |  |
| PPV | 0.04 | 91.0% |  | 54.7% |  |
|  | 0.5 | 99.6% |  | 97.5% |  |
| NPV | 0.04 | 99.9% |  | 99.8% |  |
|  | 0.5 | 97.1% |  | 95.6% |  |

PPV: positive predictive value NPV: negative predictive value

We next investigated the consistency between S1 IgG and S2-78 IgG (**Fig. 3a, b**). The overall consistencies for COVID-19 group and Control group are 96.3% and 96.7%, respectively. Interestingly, there are eight samples from COVID-19 patients are negative for S1 IgG but positive for S2-78 IgG, suggesting it will be of diagnostic value to combine S2-78 and S1. It is known that the immune response may correlate to some key clinical parameters, such as gender, disease severity, age and the final outcome[5,9]. To test whether the positive rate of S2-78 IgG is associated with these clinical parameters, we analyzed the detection sensitivities among subgroups, *i. e*., male vs. female, ≥60 vs. <60 for age, severe vs. non-severe cases and survivor vs. non-survivor with critical diseases. Similar to that of S1 IgG (**Fig. S2a**), no significant difference was observed in all these subgroups.

**Figure 3.**
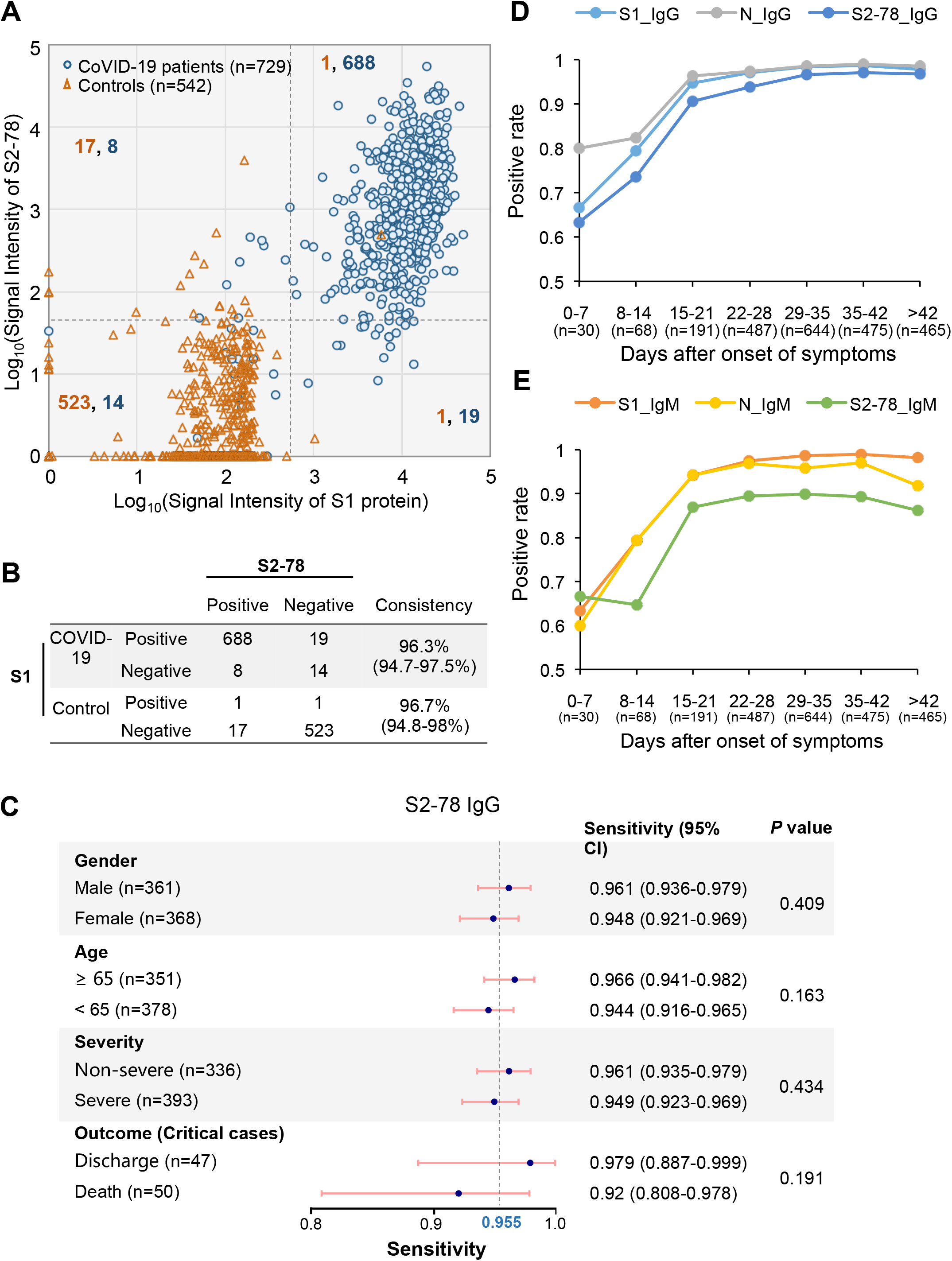
S2-78 IgG for diagnosis of COVID-19. **A**. Scatter plots of sera samples from COVID-19 (blue dots) and controls (orange triangle) for S1 IgG vs. S2-8 IgG. The grey lines indicate the cut-off values set as the optimal Youden index based on the ROC. The orange and blue numbers indicate the sample counts of control and patient on each quadrant, respectively. **B**. consistency between S1 protein and S2-78. The consistency values are provided with 95% CI. **C**. forest plot of sensitivities of S2-78 IgG among subgroups, i.e., age, gender, severity and outcome. The dots indicate the sensitivities while the error bars indicate the 95% CI. The exact values are also provided. P values were calculated with χ2 test. **D-E**. Graph of positive rates of IgG or IgM against the S, N proteins or S2-78 versus days after symptom onset in 2,360 serum samples from 784 patients.

Virus specific antibodies would persistently increase and usually conduct seroconversion within one or two weeks after symptom onset, and the portion of patients with positive antibodies reach ~100% after two or three weeks[21,22]. To test whether S2-78 IgG is suitable to detect the dynamic change of antibody response, we analyzed the positive rates at different time points after the onset of symptoms. Expectedly, the positive rate of S2-78 IgG continuously increased and reach the plateau about three weeks after onset, which is similar as S1 and N IgG (**Fig. 3d**). Similar trends were observed for S2-78 IgM (**Fig. 3e**) and IgG antibodies against other peptides (**Fig. S2b**). These observations suggest that the antibodies against S2-78 and other peptides could be applied for monitoring virus specific antibody dynamics.

### The diagnostic performance of S2-78 IgG is comparable to that of S1 IgG for asymptomatic infections

Monitoring of asymptomatic infections is essential to put SARS-CoV-2 infection under control. It is thought that asymptomatic infection usually has a weak immune response[23]. To test whether S2-78 IgG could be used for detection of asymptomatic infection, we analyzed 63 asymptomatic patients (Cohort 4, **Table 1**) defined as positive either in NAT test or antibody test conducted by a commercial assay (see **methods**) but without obvious symptom. Four subgroups were divided according to positive or not for NAT, IgG or IgM[24]. It was shown that for S1 IgG and S2-78 IgG, all samples (n=4) of IgG^-^ group were negative, while of the 59 IgG^+^ asymptomatic infections, 47 and 45 were positive for S1 IgG and S2-78 IgG, respectively. (**Fig 4a, b**). The consistency between S1 IgG and S2-78 IgG was also high (93.7%). The contradictory results in IgG^+^ group between our data and the commercial assay may due to the differences of antigens involved. S and N recombinant proteins are used for the commercial assay, for which slightly lower specificity is common[7]. Overall, these results demonstrate the diagnostic and screening value of S2-78 IgG for asymptomatic infections.

**Figure 4.**
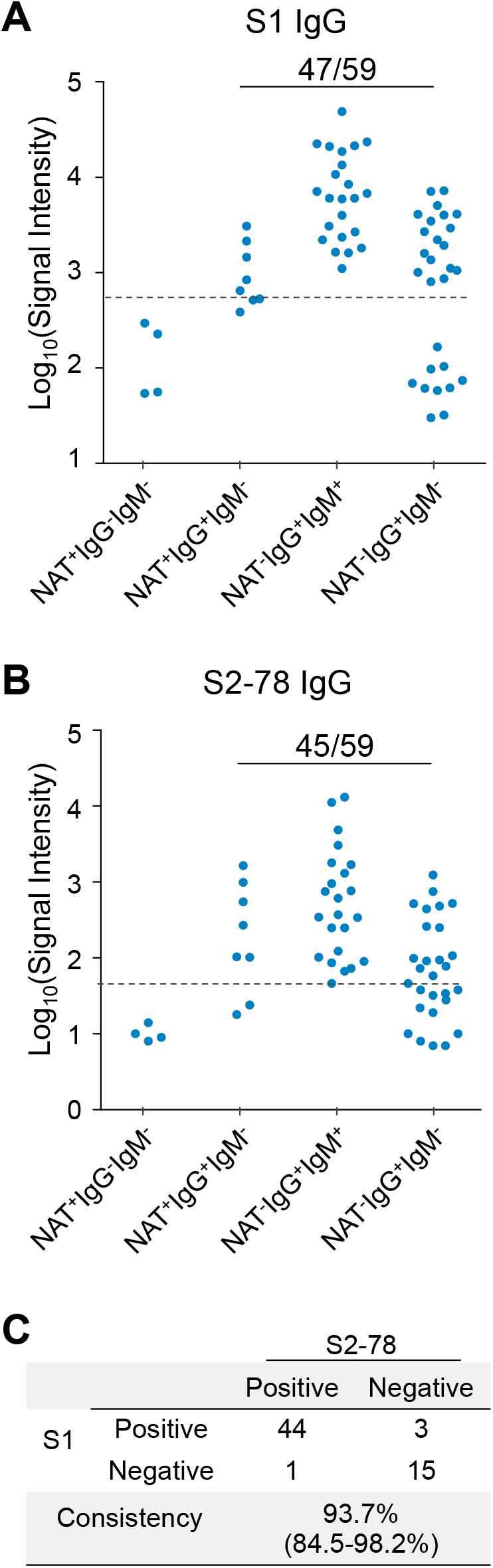
S2-78 IgG for diagnosis of asymptomatic patients. **A-B**. Levels of S1 IgG and S2-78 IgG in asymptomatic patient groups divided by positive or negative among three tests, i.e., NAT (nucleic acid test), SARS-CoV-2 vims specific IgG and IgM antibodies. The dashed lines indicate the cut-off values. The number of positive samples as well as the total number of samples of IgG^+^ (IgG positive) groups were presented for S1 IgG and S2-78 IgG. **C**. The consistency between S1 IgG and S2-78 IgG.

### The diagnostic value of S2-78 IgG was validated by ELISA

ELISA (enzyme linked immunosorbent assay) is common for commercial SARS-CoV-2 antibody assays[8,25]. To verify the efficacy and applicability of S2-78 IgG, we established an ELISA assay. Firstly, to test the consistency of the peptide microarray and ELISA, we randomly selected 31 sera from COVID-19 patients of Cohort 1 and tested by ELISA. High consistency was achieved with a Pearson correlation of 0.926 (**Fig. 5a**), demonstrating the validity of the microarray results. To further validate the performance of S2-78 IgG, we screened another independent cohort of samples collected from a different medical center (Cohort 3, **Table 1**). As expected, high performance of S2-78 IgG for specific detection of COVID-19 was achieved by ELISA (**Fig. 5b, c**).

**Figure 5.**
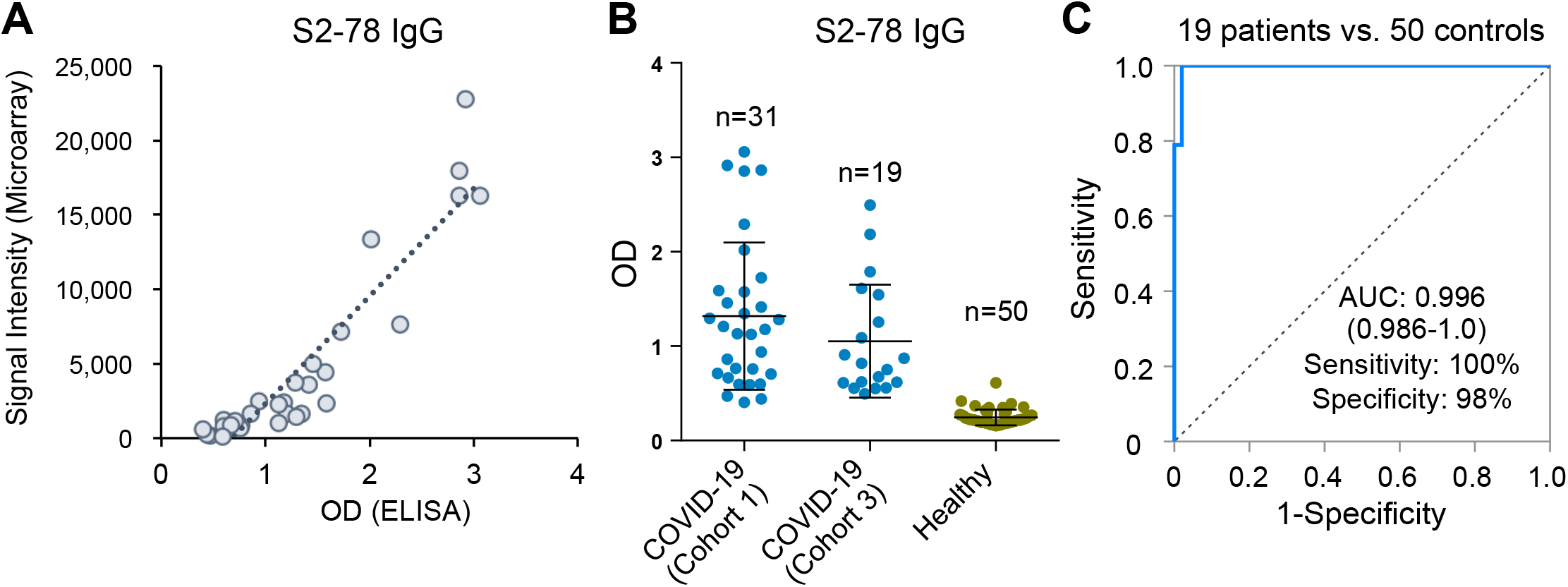
ELISA validation of S2-78 by Cohort 3. **A**. Correlation of the signals of S2-78 IgG response between peptide microarray and ELISA with Cohort 1 (n=31). **B**. Levels of S2-78 IgG in samples from COVID-19 patients either from Cohort 1 (n=31) or Cohort 3 (n=19) and healthy controls (n=50). **C**. ROC of S2-78 IgG for discrimination of 19 COVID-patients (Cohort 3) and 50 healthy controls.

### A two-step strategy by combining S2-78 and other peptides for specific detection of SARS-CoV-2 infections

Because of the highly homologous genomes among human coronaviruses, one unneglectable risk for antibody based diagnosis of SARS-CoV-2 infection is the possible cross-reactivity with other coronaviruses, especially for those common cold causing coronaviruses, *i. e*., HCoV-OC43, HKU1, NL63 and 229E. The protein sequences of S protein among those viruses are of high similarity, selection of sections of S protein that are unique for SARS-CoV-2 is important for diagnosis[11,13]. To investigate whether the identified peptides are specific to SARS-CoV-2, we performed homology analysis among SARS-CoV-2 and 6 other coronaviruses (**Extended data Fig 3**). High homologies were observed for S2-78 and S2-22, suggesting they alone may not suitable for specific detection of SARS-Cov-2 infection. In contrast, other peptides, *i. e*, S1-93, 97,101,105,111 and S2-97, exhibit low similarities with other coronaviruses, particularly the four coronaviruses that cause common cold in human, suggesting they could be applied for specifically detection of SARS-Cov-2 infection. S1-97 with the best performance among these peptides was selected for further investigation. Considering the relatively low sensitivity (86.2%) of S1-97 IgG (**Fig. 2, Fig. S4**), we proposed a two-step strategy by combining S1-97 IgG and S2-78 IgG for detection (**Fig. 6a**). To simplify this strategy, there are three assumptions: 1) S2-78 IgG can detect the infection of SARS-CoV-2 and related coronaviruses with the same sensitivity (95.5%) and generates a false positive rate of 3.3% (1-speficicity, 1-96.7%) for healthy and patients of other diseases; 2) S1-97 IgG can specially detect SARS-CoV-2 infection with 86.2% sensitivity and no cross-reactivity with related coronaviruses, so it would generate a false positive rate of 6.8% (1-speficicity, 1-93.2%) for the infection of related coronaviruses, healthy control and patients with other diseases; 3) The positive rate of S2-78 IgG and S1-97 IgG in each group is independent to each other. According to the two-step strategy (**Fig. 6a**), for any given sample, the first step is to detect S2-78 IgG; For the positive ones, the second step is to detect S1-97 IgG. The samples positive for both S2-78 and S1-97 are defined as final positive, while the samples negative for either one is defined as negative. As a result, the sensitivity for SARS-CoV-2 infection detection would be 82.3%, the specificity for related coronavirus infections would be 93.5% *(specificity* 1), and the specificity for control group would be 99.8% *(specificity* 2). To further improve the performance of the two-step strategy, a panel (Panel-A) of peptides was composed by bivariate regression analysis based on the specificity of each peptide to SARS-CoV-2. The linear function for Panel-A is y =0.014* *x*_1_ + 0.02* *x*_2_ - 0.003* *x*_3_ + 0.006* *x*_4_-2.593, where *y* represents signals of Panel-A and *x*_1_*, x*_3_*, x*_3_, *x*_4_ represents the signals of IgG against S1-93, 97, 101 and 105, respectively. The sensitivity and specificity of Panel A, based on the data generated from Cohort 2, is 88.3% and 96.7%, respectively (**Fig. 6b, c**). Follow the two-way strategy by combining S2-78 IgG and Panel-A, the final sensitivity for the detection of SARS-CoV-2 infection, specificity for related coronavirus infection *(specificity* 1), and the specificity for control group *(specificity 2)* are 84.3%, 96.8% and 99.9%, respectively (**Fig. 6d**). When S1 protein and Panel-A are combined, the final sensitivity for the detection of SARS-CoV-2 infection, specificity for related coronavirus infection *(specificity* 1), and the specificity for control group *(specificity 2)* are 85.3%, 96.8% and 99.99%, respectively (**Fig. S4b**). These results demonstrate that combination of human coronavirus conserved peptides or S1 protein and other SARS-CoV-2 specific peptides (“significant” peptides) enable specific detection of SARS-CoV-2 infection with a high specificity and acceptable sensitivity.

**Figure 6.**
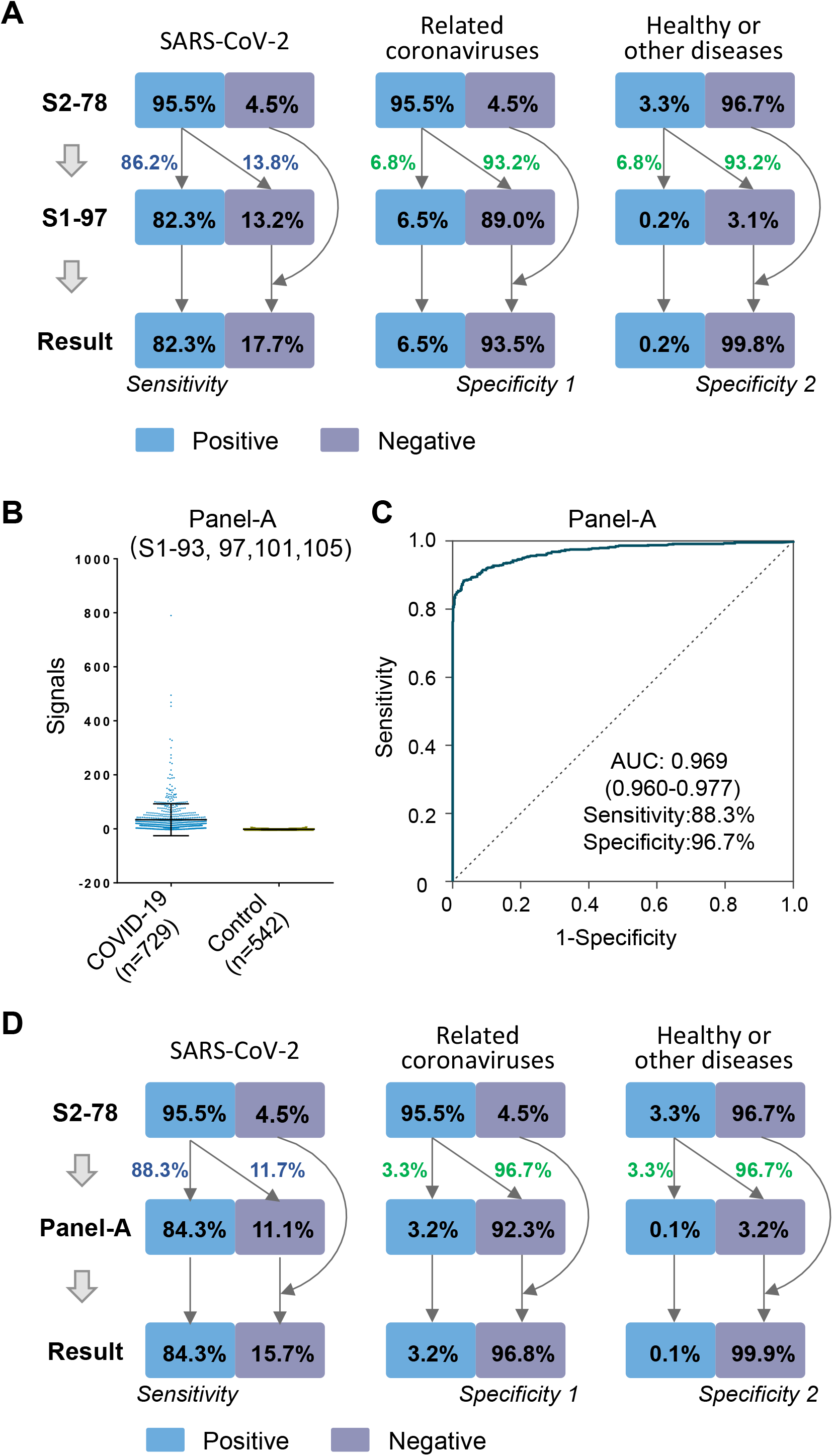
Discrimination of related coronaviruses with combination of peptides. **A**. Illustration of the strategy with combination of S2-78 IgG and S1-97 IgG based on the results of Cohort 2 (729 patients and 542 controls). For any given sample, the first step is to detect S2-78 IgG response. For the positive ones, the second step is to detect S1-97 IgG response. The blue numbers indicate the positive or negative rates for SARS-CoV-2 infections, while the green numbers indicated the assumed positive or negative rates for samples not belong the SARS-CoV-2 group. The last step summaries the overall sensitivities and specificities for different sample groups. **B**. Signals of Panel-A (S1-93, 97, 101 and 105 in COVID-19 patients and control groups. **C**. The corresponding ROC of Panel-A. **D**. The performance of the combination of S2-78 IgG and Panel-A.

## Discussion

In this study, we took advantage of a peptide microarray of full S protein coverage [17,26], analyzed 2,434 sera from 858 COVID-19 patients, sera from 63 asymptomatic patients and 610 controls collected from multiple medical centers. We identified eight 12-mer peptides (“significant” peptides) which exhibit high diagnostic values as antigens to detect SARS-CoV-2 specific IgG or IgM. Among the “significant” peptides, S2-78 IgG has a comparable diagnosis performance to that of S1 protein for the detection of COVID-19 and asymptomatic infections, suggesting S2-78 has the potential for serological prevalence investigation. By combining S2-78 and other “significant” peptides of low homologous among common human coronaviruses, we purposed a two-step strategy for accurate and affordable detection of SARS-CoV-2 infection.

Serological test plays an important role in diagnosis and monitoring of COVID-19. Recombinant proteins, particular S1, is one of the key reagents to build immunoassay for detecting SARS-CoV-2 IgG, IgM or IgA. However, expressing S1 protein in the right conformation is usually difficult, and in some cases, the antibodies that recognize membrane spike protein are unable to bind recombinant S protein[27]. Moreover, the inconsistency of the proteins from different manufacturers and even different batches from the same manufacturer may result in high variation. High cost and insufficient capacity to produce enough amount of high quality recombinant S1 protein limits the accessibility of the immunoassay in poor or remote regions around the world. Alternatively, peptide-based immunoassay provides a superior choice to S1 protein assays. The reasons are as follows: 1) The peptide synthesis could be easily scaled up when required. A large amount of peptide could be easily synthesized within a very short period of time, if necessary, the peptide could be synthesized in a GMP facility; 2) The consistency and purity of peptide synthesis is high, there is almost no batch-to-batch variation; 3) Peptide is very stable, as reagent, it could be easily stored and transported; 4) The cost of peptide is about 2-3 magnitude lower than that of S1 protein.

We identified S2-78 as a candidate of high diagnostic value. Applying large cohorts of COVID-19 patients and a variety types of controls, we comprehensively verified S2-78 IgG as a good candidate for diagnosis of COVID-19, with comparable specificity and sensitivity to that of S1 IgG. Expectedly, we found that the overall consistency of S2-78 IgG and S1 IgG is high, suggesting S2-78 has the potential to replace S1 protein. It is notable that, the signal intensity level of S2-78 IgG is lower than that of S1 IgG, which is reasonable since there are multiple sites on S1 protein that could be recognized by antibodies in COVID-19 sera. However, because it is hard to ensure extremely high purity when purifying recombinant proteins, the background of S1 IgG for control group is also higher than that of peptides. These results imply the sensitivity of S2-78 IgG might be further evaluated when raise the antigen concentration or adopt a more sensitive platform for detection, such as electrochemical platform or single-molecule detection technologies[28,29]. Indeed, we also performed S1 based ELISA assay by a commercial kit with the same set of samples, the overall performance of S2-78 IgG based ELISA is comparable to that of the commercial kits (data not shown).

Immunoassay is the major tool to assess the extent of virus circulation in population and the likelihood of protection against re-infection by screening population to identify infected individuals without clinical symptoms[7,12,19]. We verified that S2-78 IgG could be applied to detect asymptomatic infections with a comparable sensitivity and specificity with S1 IgG, though these individuals are thought to have weaker immune responses[23]. We assessed PPV and NPV for two prevalence rates, *i. e*., 0.04 and 0.5. For 0.04, the PPV is 54.7% and NPV is 99.8%, indicating a high false positive rate, while it can effectively and accurately exclude the negative ones. Due to the small number of positive cases under this circumstance, one possible solution is to re-test the suspected samples by additional assays. Moreover, to meet the requirement of population-wide application, the performance of S2-78 might be improved by optimizing the parameters or adoption of other platforms.

S protein shares high sequence similarities with other seasonal circulated human coronaviruses. Theoretically, cross-reactivity may exist when S/S1 is applied as antigen for immunological test, thus cause false positive. It is thus necessary to pinpoint specific regions/ sites of SARS-CoV-2 S protein and eliminate the potential cross-reactivity. Selection of peptides with high antibody responses and low sequence similarities with other coronaviruses will improve the performance of diagnosis. Among the identified peptides, except S2-78 and S2-22, other peptides are very distinct to the four circulating human coronaviruses, implying that they could be served as the specific antigen to eliminate the potential cross-activity. However, these peptides exhibit slightly lower sensitivity and specificity. To take advantage of these “significant” peptides, we proposed a two-step strategy that combine S2-78 with other significant peptide/s to discriminate SARS-CoV-2 from related infections as well as non-infections. This study could be further strengthened by testing sera collected from the infections of common human coronaviruses, *i. e*, HCoV-OC43, HKU1, NL63 and 229E.

In summary, we identified and verified eight peptides derived from S protein that exhibit high diagnostic values. These peptides might be used in different circumstances alone or in combination as candidates to build immunoassay/s for monitoring COVID-19. In comparison to the current protein based immunoassays, the peptide based assays will be highly affordable and accessible.

## Materials and Methods

### Patients and samples

The study was approved by the Ethical Committee of Tongji Hospital, Tongji Medical College, Huazhong University of Science and Technology, Wuhan, China (ITJ-C20200128), Institutional Ethics Review Committee of Foshan Fourth Hospital, Foshan, China (202005) and the Ethical Committee of The Fifth Affiliated Hospital of Sun Yat-sen University, Zhuhai, China (K14-2). Written informed consent was obtained from all participants enrolled in this study. COVID-19 patients were hospitalized and received treatment in multiple medical center during the period from 25 January 2020 and 28 April 2020. Sera of the control group from healthy donors, lung cancer patients, patients with autoimmune diseases were collected from Ruijin Hospital, Shanghai, China or Tongren Hospital, Shanghai, China. The negative reference samples were from National Institutes for Food and Drug Control. Sera of 63 asymptomatic patients were also from Tongji Hospital, Wuhan. The IgM and IgG antibodies against recombinant nucleoprotein and spike protein of SARS-CoV-2 in sera of these patients were detected by a commercial kit (YHLO Biotech, Shenzhen, China). According to the instruction of the kit, the antibody level ≥10 AU/ mL is positive, and <10 AU/mL is negative. All the samples were stored at −80°C until use.

### Peptide synthesis and conjugation with BSA

The N-terminal amidated peptides were synthesized by GL Biochem, Ltd. (Shanghai, China). Each peptide was individually conjugated with BSA using Sulfo-SMCC (Thermo Fisher Scientific, MA, USA) according to the manufacture’s instruction. Briefly, BSA was activated by Sulfo-SMCC in a molar ratio of 1: 30, followed by dialysis in PBS buffer. The peptide with cysteine was added in a w/w ratio of 1:1 and incubated for 2 h, followed by dialysis in PBS to remove free peptides. A few conjugates were randomly selected for examination by SDS-PAGE. For the conjugates of biotin-BSA-peptide, before conjugation, BSA was labelled with biotin by using NHS-LC-Biotin reagent (Thermo Fisher Scientific, MA, USA) with a molar ratio of 1: 5, and then activated by Sulfo-SMCC.

### Peptide microarray fabrication

The peptide-BSA conjugates as well as S1 protein, RBD protein and N protein of SARS-CoV-2, along with the negative (BSA) and positive controls (anti-Human IgG and IgM antibody), were printed in triplicate on PATH substrate slide (Grace Bio-Labs, Oregon, USA) to generate identical arrays in a 1 x 7 (for the peptide microarray with three concentrations) or 2 x 7 subarray format (for the peptide microarray with one concentration) using Super Marathon printer (Arrayjet, UK). The microarrays were stored at −80°C until use.

### Microarray-based serum analysis

A 7 or 14-chamber rubber gasket was mounted onto each slide to create individual chambers for the 7 or 14 identical subarrays. The microarray was used for serum profiling as described previously with minor modifications[30]. Briefly, the arrays stored at −80°C were warmed to room temperature and then incubated in blocking buffer (3% BSA in 1×PBS buffer with 0.1% Tween 20) for 3 h. A total of 400 μL for 7-subarray format or 200 μL 14-subarray format of diluted sera or antibodies was incubated with each subarray for 2 h. The sera were diluted at 1:200 for most samples and for competition experiment, free peptides were added at a concentration of 0.25 mg/mL. For the enriched antibodies, 0.1-0.5 p,g antibodies were included in 200 μL incubation buffer. The arrays were washed with 1×PBST and bound antibodies were detected by incubating with Cy3-conjugated goat anti-human IgG and Alexa Fluor 647-conjugated donkey anti-human IgM (Jackson ImmunoResearch, PA, USA), which were diluted for 1: 1,000 in 1×PBST. The incubation was carried out at room temperature for 1 h. The microarrays were then washed with 1×PBST and dried by centrifugation at room temperature and scanned by LuxScan 10K-A (CapitalBio Corporation, Beijing, China) with the parameters set as 95% laser power/ PMT 550 and 95% laser power/ PMT 480 for IgM and IgG, respectively. The fluorescent intensity was extracted by GenePix Pro 6.0 software (Molecular Devices, CA, USA).

### Data analysis of peptide microarray

For each spot, signal intensity was defined as the foreground subtracted by the background. The signal intensities of the triplicate spots for each peptide or protein were averaged. For the samples of Cohort 2 and 3, normalization among microarray slides were performed. For each slide, block #14 was incubated with the positive reference sample which was generated by pooling of 50 randomly selected sera from COVID-19 patients. For each slide, the data generated from the positive reference sample were subjected to build a linear regression function, and the data from other samples were subjected to linear normalization according to the function. Graphpad 6.0 was used to generate ROC plots and calculated AUC values.

### ELISA

Briefly, 96-well microplates with high binding polystyrene surface (Corning, New York, USA) were coated with 100 μL BSA conjugated peptide (S2-78) at 100 μg/mL and incubated overnight at 4°C. The plates were washed once with PBST buffer (PBS buffer with 0.1% Tween 20), and blocked with 3% BSA (bovine serum albumin) for 1 h at room temperature, followed by one wash with PBST. The sera were diluted at 1:50 in PBST buffer with 1% BSA, 1% FBS and 3% horse serum, 100 μL of the preparation was loaded to each well, the incubation was carried out at 37°C for 1.5 h in 100 μL. After six washes with PBST, the secondary antibody, *i.e*., anti-human IgG-peroxidase (Sangon Biotech, Shanghai, China) was diluted at 1:10000 and incubated at 37 ° C for 1 h, followed by eight washes. Tetramethylbenzidine substrate (Sigma-Aldrich, Missouri, USA) was then added and incubated for 20 min. Finally, 50 μL sulfuric acid (2 M) was added to stop the reaction. The optical density was read at 450 nm using a Behring EL311 ELISA microplate reader (Dade Behring Marburg Gmbh, Berlin, Germany). The assays were repeated twice for each sample.

## Data Availability

The peptide microarray data generated in this study has been deposited to Protein Microarray Database (http://www.proteinmicroarray.cn) under the accession number PMDE242 and PMDE244 Additional data related to this study may be requested from the authors.

**Abbreviations**
AID: autoimmune diseases
AUC: area under curve
CI: confidence interval
CoV: coronavirus
ELISA: enzyme linked immunosorbent assay
GMP: good manufacturing practice
NAT: nucleic acid test
NPV: negative predictive value
PPV: positive predictive value
ROC: receiver operating characteristic
SARS: severe acute respiratory syndrome
URI: upper respiratory infections
WHO: world health organization.

## Acknowledgments

We thank Dr. Daniel M. Czajkowsky for critical reading and editing.

## Author contributions

S-C.T. developed the conceptual ideas and designed the study. Z-Y.S., F.W., H-Y.H., Y-D.Z., X-S. L., Z-J.Y., H-M.S., J-X.W., L-Y.C., S-Q.L., P-F.P., and H.S. collected the sera samples. X-L.F., Y.L., D-Y.L., Q.L., Z-W.X., M-L. M., B.Z., H.C., C-Z.Y., J-B.X., X-N.W., Y-X.Z., H-N.Z., H-W.J., H.Q. Y-D.Z., X-S.L., Z-J.Y and S-J.G. performed the experiments and analysis. S-C.T. and Y.L. wrote the manuscript with suggestions from other authors.

---

**Figure S1.**
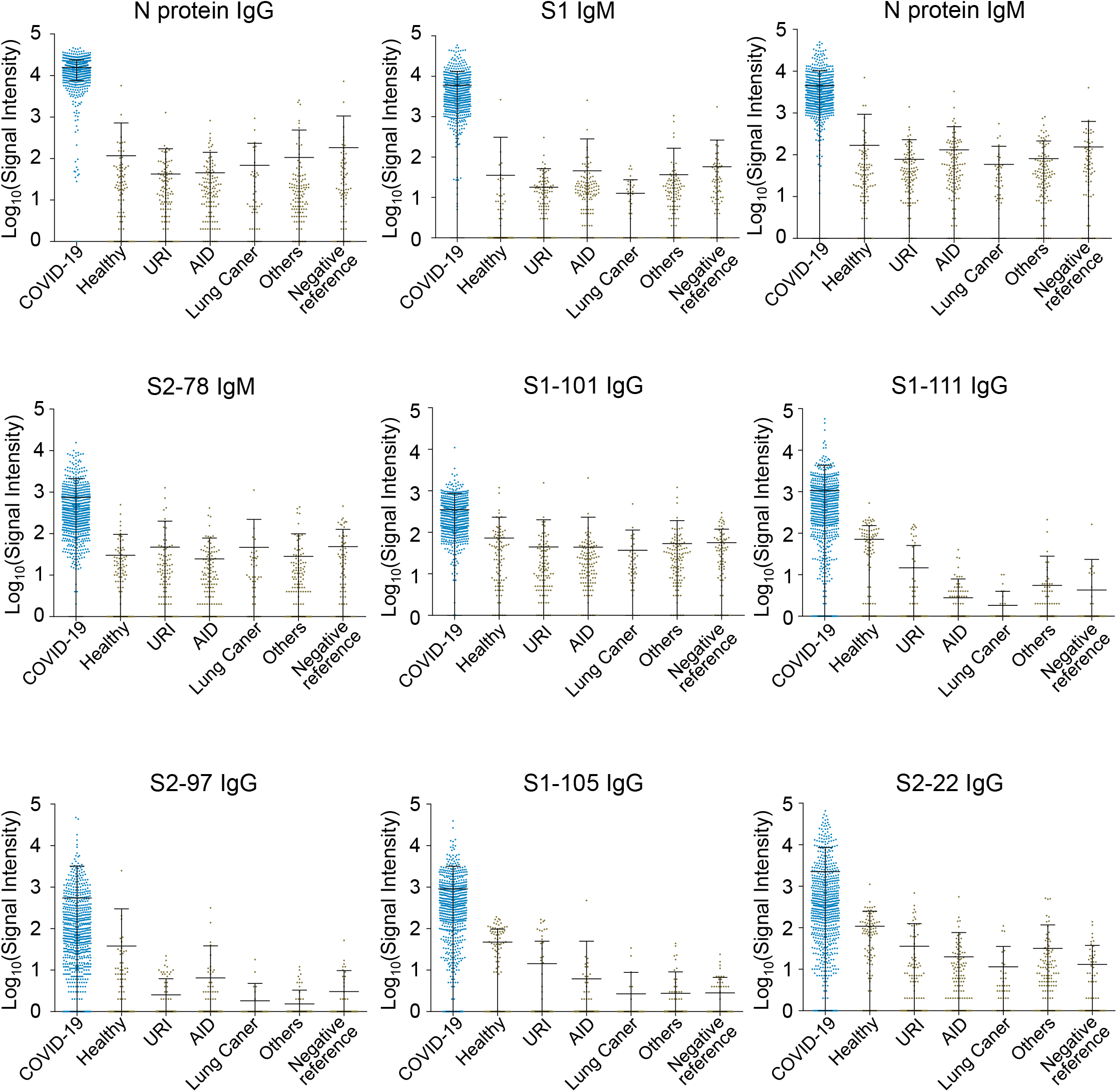
Evaluation of the significant peptides by Cohort 2 - other peptides. Signal levels of the antibodies against the indicated peptides in COVID-19 patients (n=729), Healthy controls (n=92), upper respiratory infections (URI, n=104), patients with autoimmune diseases (AID, n=120), lung cancer patients (n=41), patients with other diseases (n=112) and negative reference samples (n=73) of Cohort 2.

**Figure S2.**
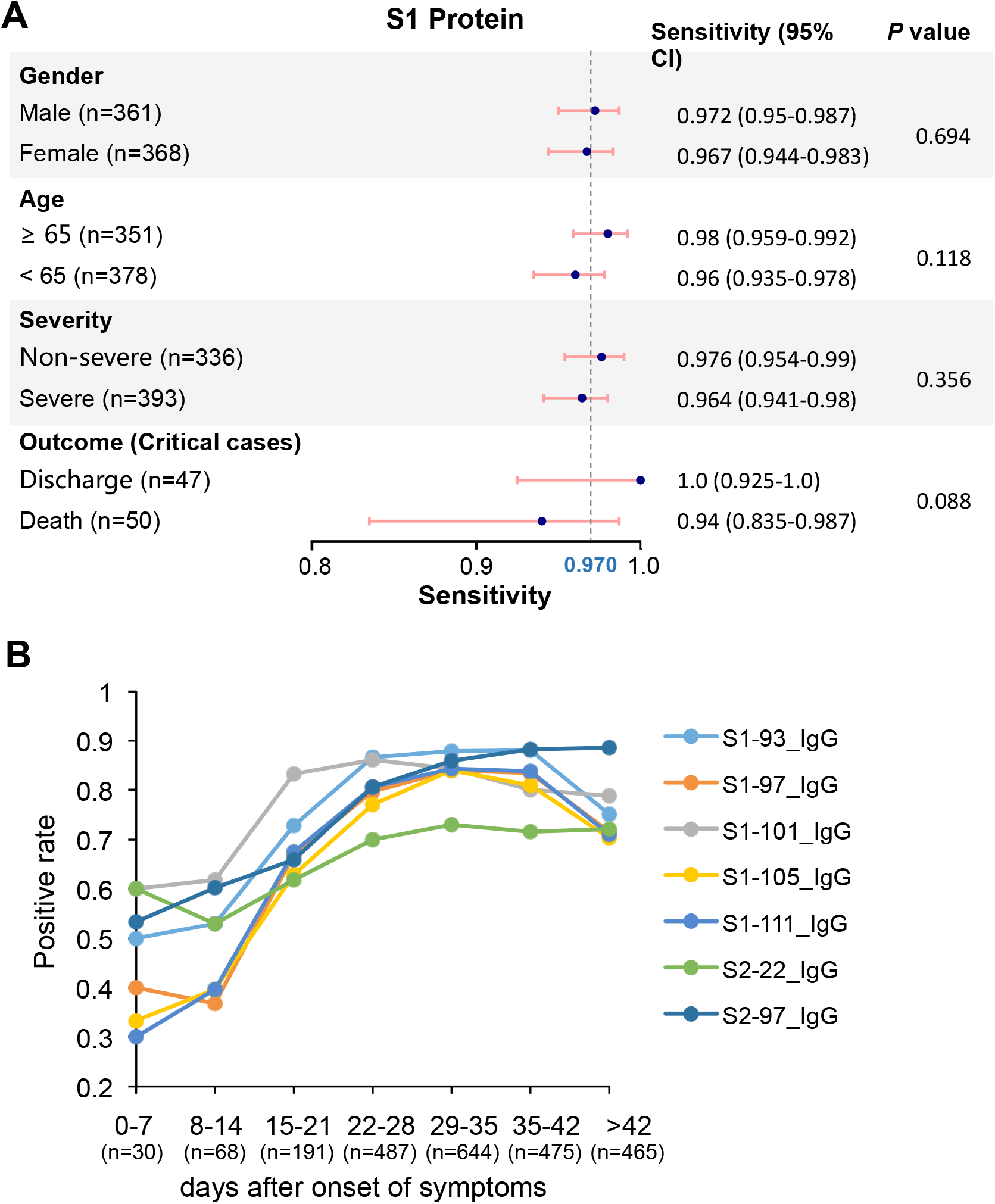
Performance of peptides and S1 for diagnosis in subgroups. **A**. forest plot of sensitivities of S1 IgG in different subgroups, i.e., age, gender, severity and outcome. The dots indicate the sensitivities while the error bars indicate the 95% CI. The exact values are also provided. P values were calculated with χ2 test. **B**. Graph of positive rates of IgG antibodies against the indicated peptides versus days after symptom onset in 2,360 serum samples from 784 patients.

**Figure S3.**
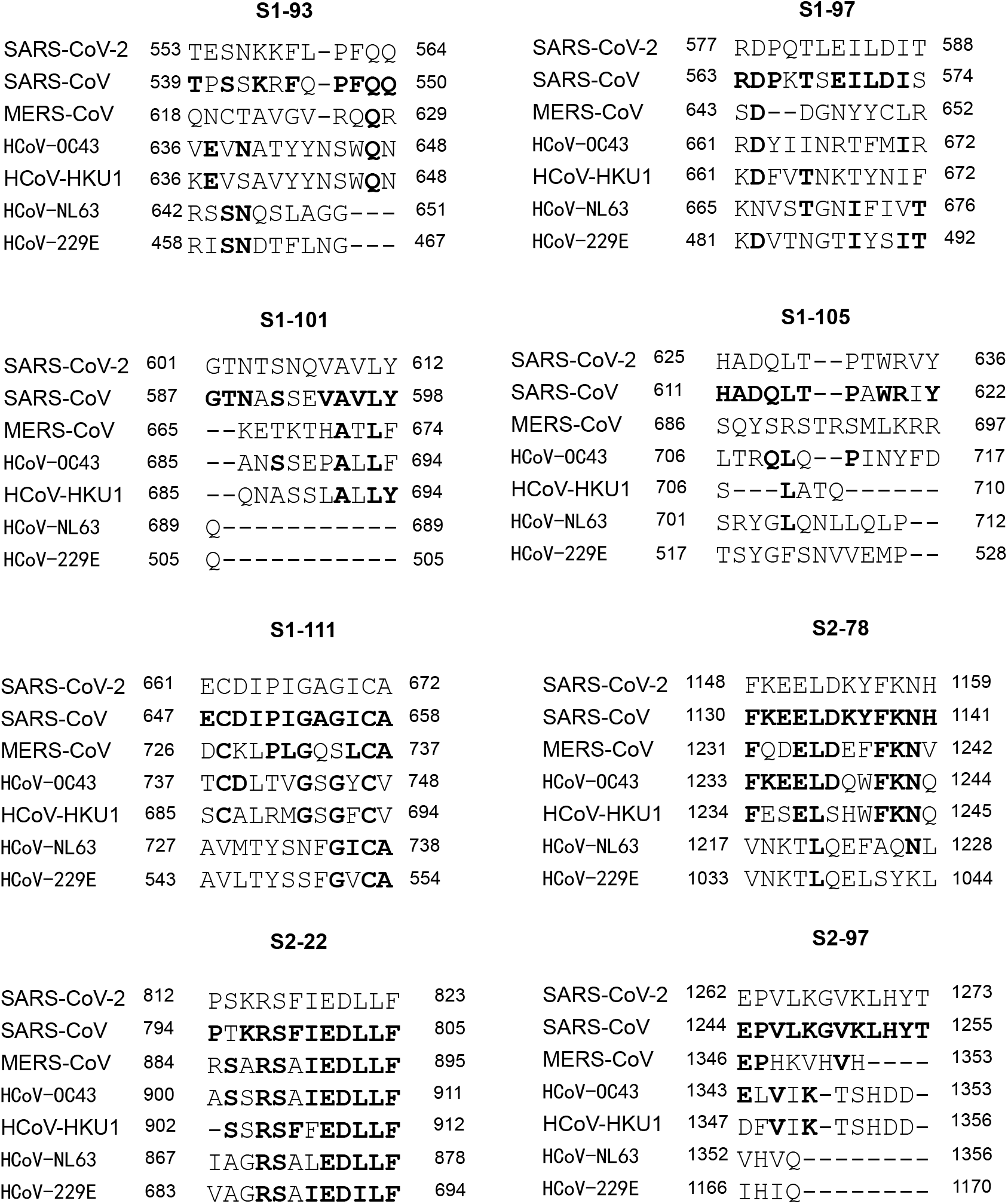
Homology analysis of the significant peptides among SARS-CoV-2, SARS-CoV, MERS-CoV and other four human coronaviruses. The line for each position indicates the gap. The bold letters indicate the positions with the same amino acids as that of SARS-CoV-2. The homology analysis was performed by an online tool called Clustal Omega (https://www.ebi.ac.uk/Tools/msa/clustalo/).

**Figure S4.**
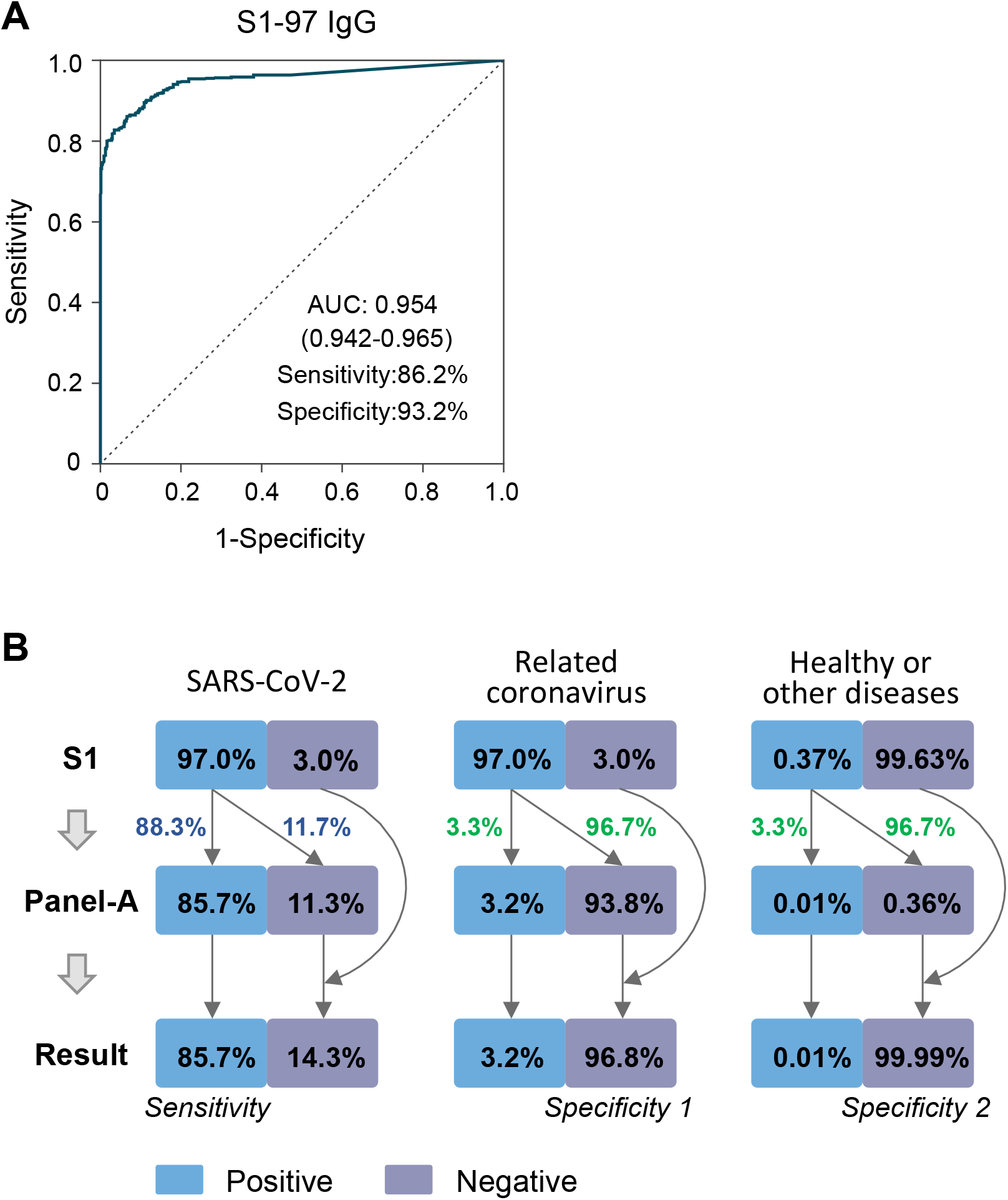
Discrimination of related coronaviruses by combining S1 and peptides of Panel-A. **A.** ROC of S1-97 IgG based on the results of Cohort 2. **B**. The combination of S1 IgG and Panel-A based on the results of Cohort 2 (729 patients and 542 controls).

**Table S1.**
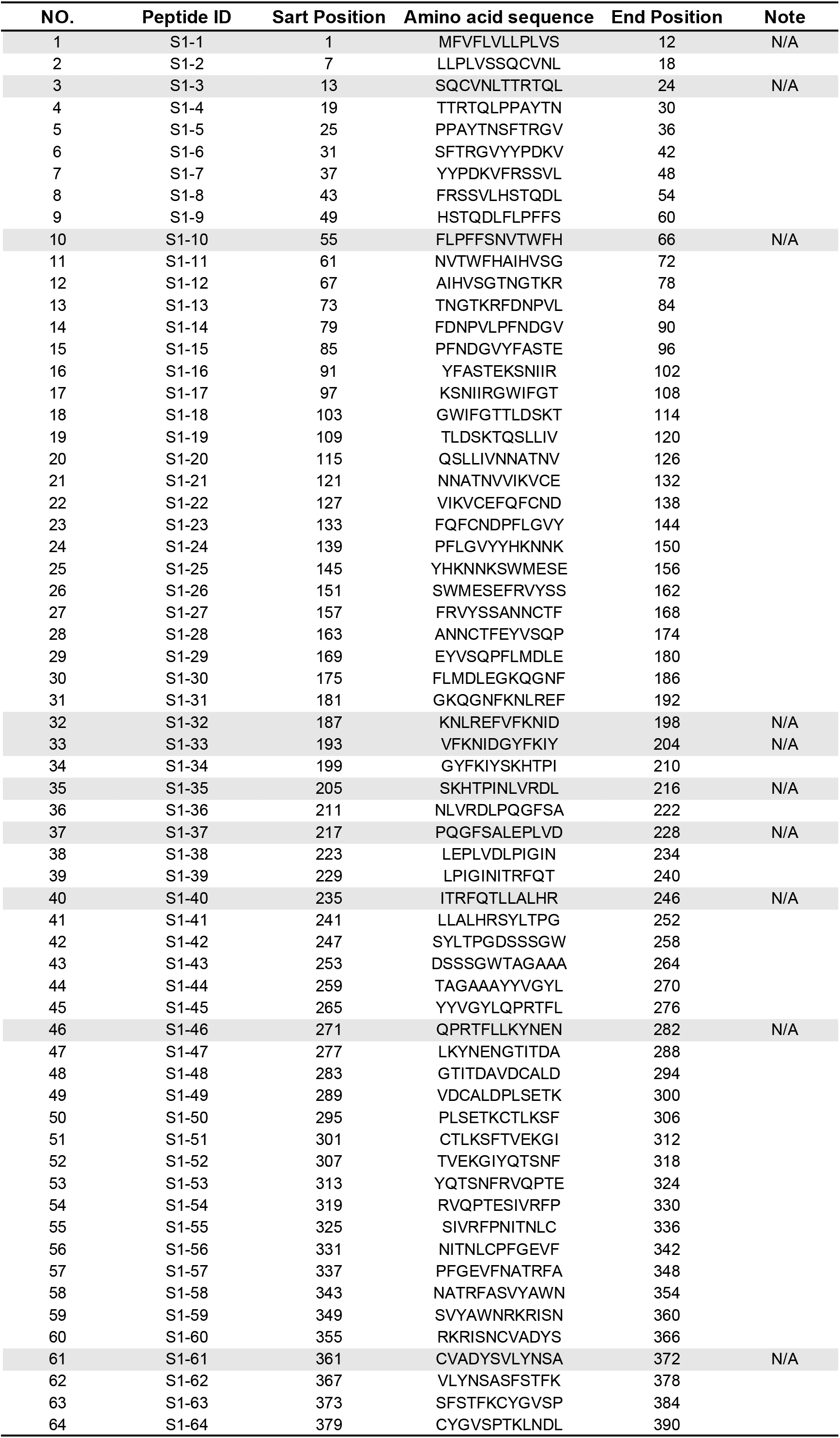
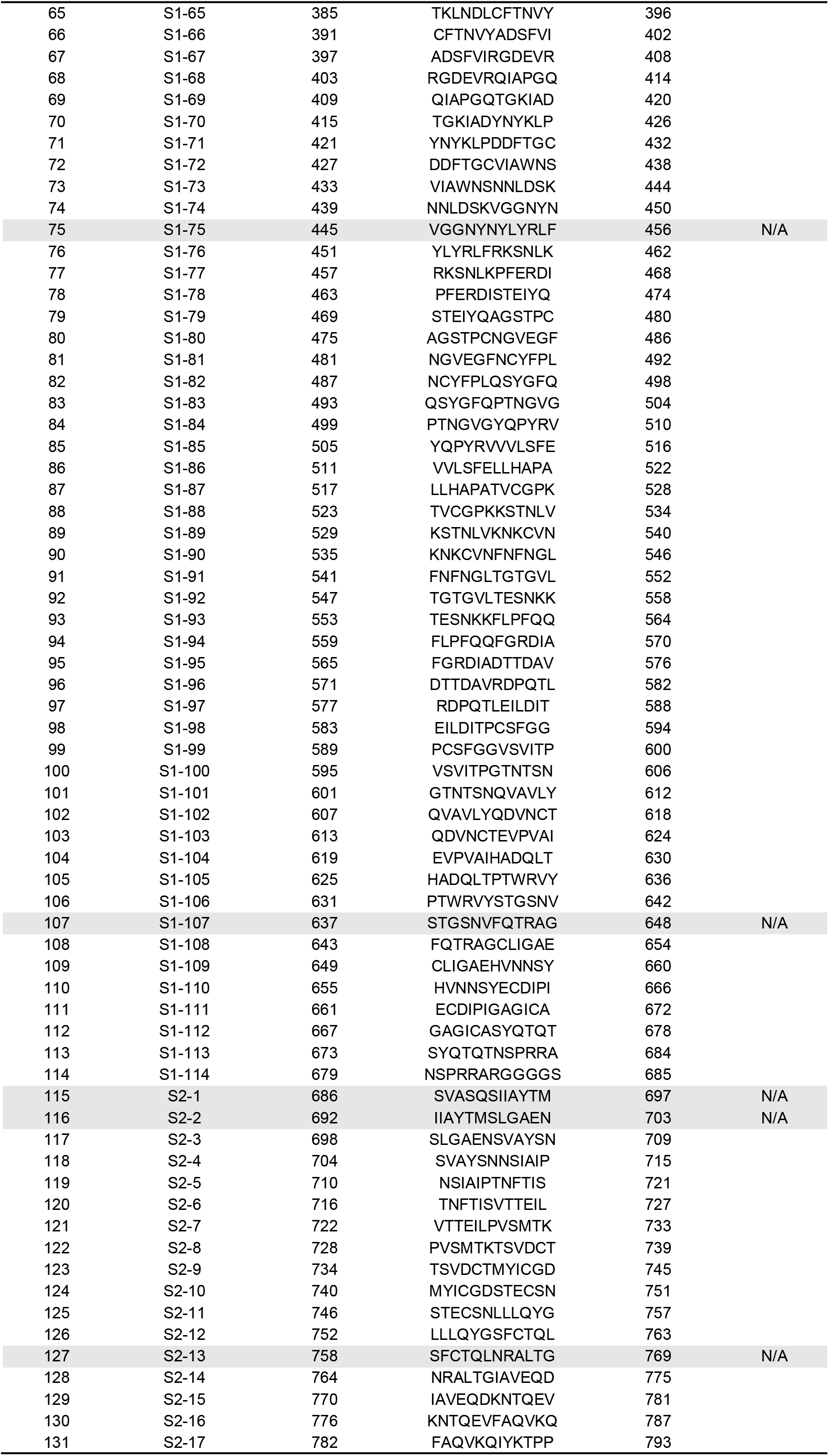
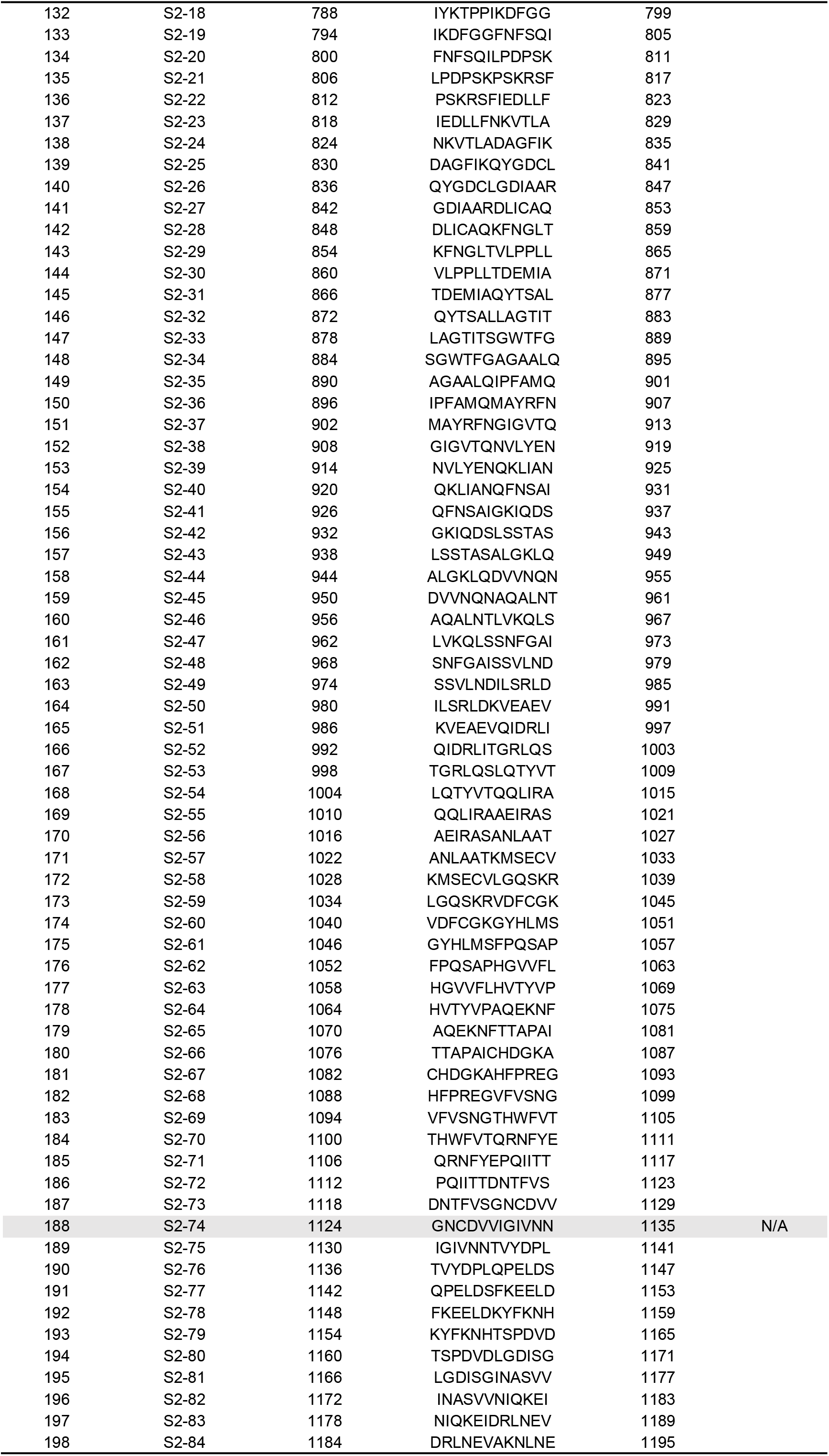
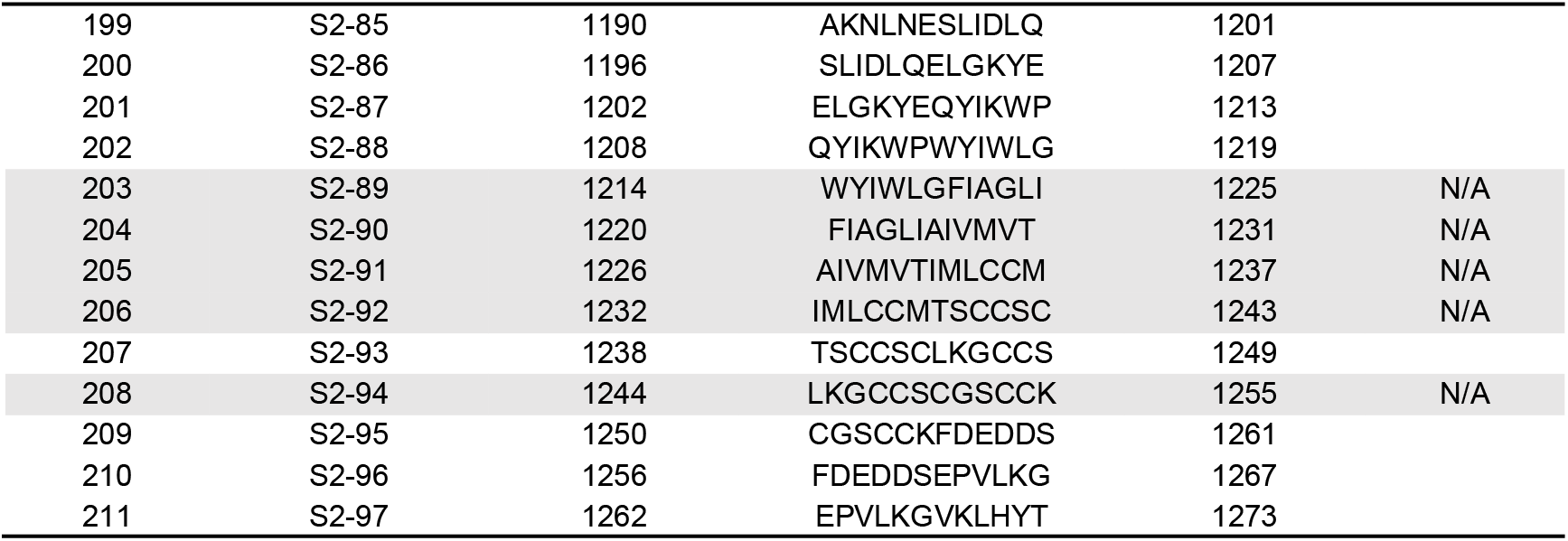
The peptides synthesized in this study.

